# Longterm kinetics of vibriocidal antibody responses after *V. cholerae* infection in the Democratic Republic of Congo

**DOI:** 10.1101/2025.02.26.25322966

**Authors:** Kilee L. Davis, Carmen Nieznanski, Felicien Maisha, Ben J. Brintz, Christy Clutter, Meer Alam, Cyrus Saleem, Afsar Ali, J. Glenn Morris, Daniel T. Leung

## Abstract

Interpretation of sero-epidemiology studies of cholera relies on knowledge of *Vibrio cholerae* antibody kinetics, which thus far have been lacking in African populations. We performed vibriocidal assays on 212 serum samples from 116 culture-positive cholera patients (median age 8 (IQR 4-18)) in Goma, DRC, collected at enrollment and again 3-449 days after. Vibriocidal responses peaked 7-40 days after symptom onset, with nearly all samples declining to a titer of 160 or lower by day 180. We used a Bayesian exponential decay model to show an 88% probability of a faster decay in children under 5.

## Introduction

Cholera is an acutely dehydrating diarrheal disease caused by the aquatic Gram-negative toxigenic bacterium *Vibrio cholerae* [1]. *V. cholerae* is transmitted via the ingestion of contaminated food and/or water making it most prevalent in poverty-afflicted communities around the world where water and sanitation infrastructure is limited [2]. Surveillance for cholera can be challenging due to limited capacity for disease surveillance in places where the disease occurs, variations in its seasonality, and the non-specificity of cholera symptoms [3] Additionally, laboratory confirmation has a limited impact on treatment and is infrequently performed. An emerging method for estimating cholera burden is sero-surveillance, involving measurement of cholera-specific antibody in blood obtained from a representative sampling of a defined population. Aside from burden estimates, sero-surveillance can also be potentially used to estimate a population’s immunity level. Such knowledge can inform public health strategies, including the timing of deployment of interventions such as vaccination.

Longitudinal studies examining kinetics of antibody responses to cholera are crucial for interpreting cross-sectional sero-surveillance data. Until now, kinetics of cholera antibody response have been mostly limited to patients with severe cholera in Bangladesh (a hyper-endemic region) [4]. There is a paucity of data on the kinetics of antibody responses against cholera in endemic regions outside of South Asia. Our primary aim is to investigate the long-term kinetics of *V. cholerae* antibody responses among patients in Goma, DRC, a cholera-endemic region in Africa. Our secondary aim is to compare the antibody kinetics between young children and older persons.

## Methods

### Patient recruitment and collection of samples

To examine longitudinal antibody responses to cholera, we measured vibriocidal antibody titers of patients with culture-confirmed *V. cholerae* O1 infections. Study participants were recruited from patients with acute watery diarrhea who presented to cholera treatment centers operated by the Health Division of the Province of North Kivu in Goma, DRC from August 2020 to October 2023. Written informed consent was obtained from participants or their parents/guardians for those aged 17 and younger, with assent from participants aged 7 and older. Following informed consent, clinical and epidemiological data were collected into REDCap (Research Electronic Data Capture [5]) through the University of Florida Clinical and Translational Science Institute. This study was approved by the IRB of the University of Florida and the Comité Éthique de l’Université Libre des Pays des Grands Lacs.

Stool and venous blood samples were collected at enrollment. Stool cultures were performed to detect *V. cholerae* O1. Serum was separated and stored at -80°C for future analysis. Two follow-up visits were attempted to collect venous blood samples at approximately 4 weeks and 6 months post-enrollment. Time points were used to generally describe serum samples’ collection while the exact day since symptom onset was used in our analysis. The last recorded blood sample was collected in February 2024.

### Vibriocidal Assay

Vibriocidal antibodies are the best correlate of protection against cholera [14], and the best marker for prior infection [4]. We performed the vibriocidal assay for *V. cholerae* O1 Inaba and Ogawa serotypes as previously described [6]. A positive control, an IgG monoclonal antibody against *V. cholerae* O1 O-specific polysaccharide (gift of Dr. Jason Harris, Massachusetts General Hospital, Boston MA), was added to naïve serum at 6.24 µg/µL corresponding to a vibriocidal titer of 640. Heat-inactivated serum samples were duplicated, then serially diluted two-fold in saline from 1:10 to 1:10,240 in a 96-well microplate. Negative control (bacteria without serum) and positive control were included on each plate. *V. cholerae* O1 Ogawa (X25049) and Inaba (T19749) strains were grown in Luria Broth (LB) at 37°C for 3 hours, washed with saline, then resuspended to an optical density at 600nm (OD600) of 0.3. A Growth Indicating Solution was prepared with guinea pig complement (Sigma) diluted 1:10 and the bacterial suspension diluted 1:20 in saline. This solution was added to the samples and positive control wells. Plates were incubated at 37ºC at 50 rpm for 1 hour, then LB added and incubated at 37ºC without shaking for 2 hours. Plates were measured when the growth indicator reached OD600 of at least 0.2. Vibriocidal titers were defined as the reciprocal of the highest serum dilution resulting in approximately 50% of the mean OD600 of the Growth Control wells.
Samples below the detection limit were assigned a titer of 5, and those above the limit were assigned a titer of 20,480.

### Statistical Analysis

We examined the kinetics of vibriocidal titers over time, using the titer of the serotype matched by culture results. We used ggplot2 package in R (version 4.4.1.[7]) for data visualization.

Given the paucity of data on the long-term kinetics of cholera antibody responses in children under 5, we used vibriocidal titers from participants which had at least 2 paired samples to fit a Bayesian model using the cmdstanr package in R to estimate exponential decay parameters describing the amplitude of titers and the rate of decay. The model assumes the observed and logged titer values follow a normal distribution with mean 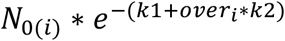 and variance *σ* where *N*_*0(i)*_ is the person-specific amplitude, *k*1 is the decay rate for the under-5 group, *over*_*i*_ is an indicator variable for being over-5, and *k*2is the change in decay rate for the over-5 group.

We fit an additional model to test for a biphasic delay with an alternative mean: 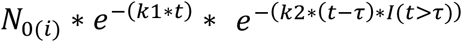 where *I*(*t* > *τ*) = 1 if *t* > *τ*, otherwise *I*(*t* > *τ*) = 0. Given that in preliminary analyses, the majority of patients’ initial vibriocidal titer was their highest titer, we assumed that we missed the initial rise in titer value to the amplitude and thus our model equation doesn’t include the rise in titer value and only examined the rate of decay.

Correspondingly, we also excluded any subjects’ titers if they had a titer below limit of detection between 3 and 60 days of follow-up, or those who never mounted a detectable antibody response. We use uninformative priors.

## Results

We performed vibriocidal assays on serum from 212 samples from 116 culture-confirmed *V. cholerae* patients (53% Male; Median age 8 (IQR 4-18)). Of the 101 participants for which serotype information was available, 83 (82%) of participants were infected with Ogawa serotype, 15 (15%) with Inaba, and 3 (3%) with Hikojima. Children under 5 represented 37% of the participants and 27 (25%) participants self-reported prior oral cholera vaccination. Twenty (20%) participants had serum samples from three time points, 57 (56%) had 2 timepoints and 38 (38%) one time point. Sample collection from many households was delayed due to unanticipated problems in reaching patient homes for repeat visits and restrictions on travel related to safety concerns. Because of this, there was substantial variability in the time of collection of what were initially scheduled as four week and six-month post-infection visits. Consequently, data were recorded and analyzed as number of days between the onset of illness and the date of sample collection, with second samples collected between 3 to 297 days, and third samples collected between 25 to 449 days after symptom onset (Fig. 1).

Overall, the highest vibriocidal responses were noted between 7 and 40 days and waned over time, with nearly all waning to a titer of 320 or lower by 90 days, and to a titer of 160 or lower by 180 days (Figure 1). We then used data from all participants which had at least 2 paired samples to examine differences in decay kinetics between participants 5 years of age and younger compared with those over 5 (Figure 2). In the exponential decay model, we found a negative posterior mean for *k*_2_, the parameter indicating a change in decay for the participants over 5, with 95% posterior credible intervals overlapping 0 (*k*_2_: -0.0012 (-0.0030, 0.0004)). The draws from the posterior distribution of *k*_2_ suggest there is an approximately 88% probability of a slower decay in participants over 5. Given that prior vaccination may increase the probability of a slower decay, we performed a sensitivity analysis among those without prior vaccination and found similar results (*k*_2_: -0.0014 (-0.0041, 0.0007), with an approximately 83% probability of a slower decay in participants over 5). In the biphasic decay model, we estimate an initial decay of *k*_1_ (0.0094 (0.0057, 0.00139)) and found evidence of a positive *k*_2_ (0.0017 (0.0007, 0.0026)) suggesting a slower decay following 62 (37, 90) days since symptom onset.

**Figure 1:**
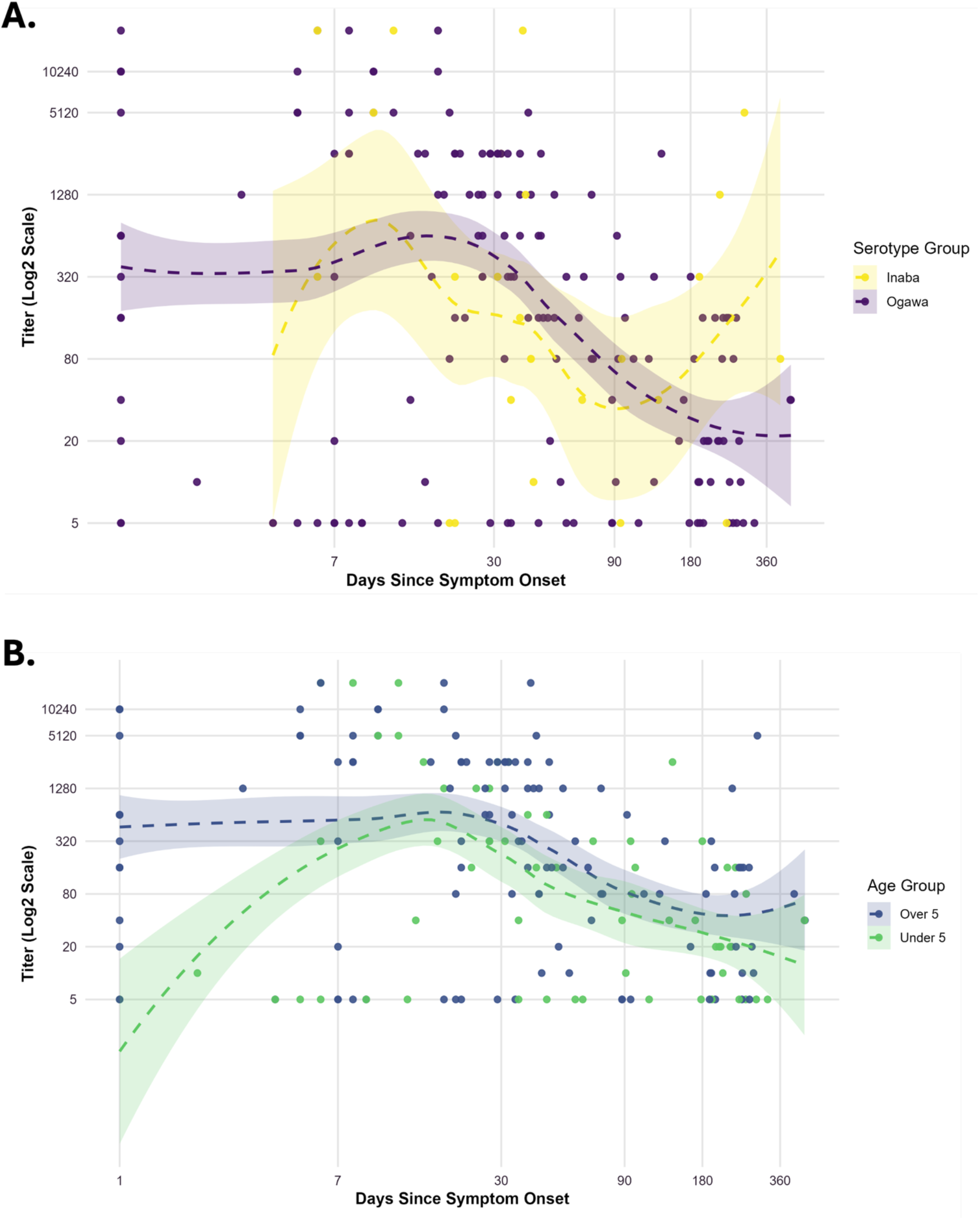
Post-infection vibriocidal titer responses from confirmed cholera cases. Vibriocidal titers by matching *V. cholerae* O1 serotype from 101 participants, by (A) serotype, and (B) age group. Days since symptom onset on the x-axis, and vibriocidal titer on the y-axis, are plotted on log2-transformed scales. Hashed lines are LOESS smoothed data with confidence intervals in shade.

**Figure 2.**
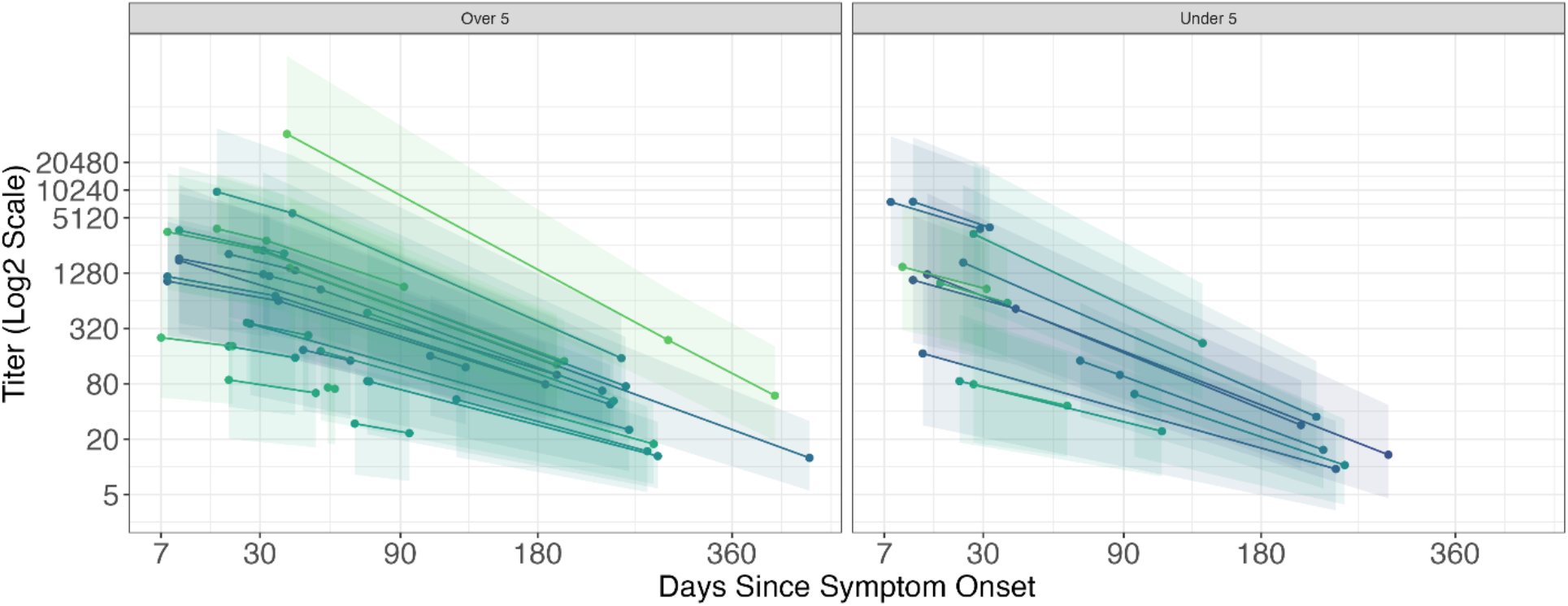
Estimated average titer value and 95% posterior credible interval for serotype-matched vibriocidal titers from the exponential decay equation plotted against days since symptom onset, by participants over 5 years of age (left) and under 5 (right). Each line represents the vibriocidal titer decay of a participant with at least 2 paired samples, fitted to a Bayesian model to estimate exponential decay parameters describing the amplitude of titers and the rate of decay.

## Discussion

In this longitudinal study of culture-confirmed cholera patients in Goma, DRC, we describe the vibriocidal antibody kinetics of patients from a cholera-endemic area in Africa, for up to one year post symptom onset. We show that vibriocidal antibody responses from children under 5 likely wanes more rapidly than those in older children and adults, and that both groups had few with titers above 160 at one-year post-infection. These data suggest that young children may be more vulnerable to repeat infection than adults, though both groups may require vaccination to boost immunity.

The predominance of young children in our study is unique compared to prior studies of vibriocidal kinetics, which have featured mostly older children and adults. Leveraging this, and the longitudinal nature of the study, we show that the waning of the vibriocidal response in young children (age 5 and lower) is likely faster than that of older persons. While the 95% posterior credible interval contains 0, a null result in the frequentist sense, our Bayesian model estimates a high (88%) probability that there is a differing decay rate between age groups. This finding fills a knowledge gap, suggesting that interventions such as the use of vaccination to boost immune response, may benefit even those in this age group who had a recent infection.

We found that the vibriocidal antibody response decays over time in a biphasic manner, results consistent with a previous study of vibriocidal kinetics in medically attended severe cholera cases in Bgnaldesh [8]. Due to differences in laboratories, assay controls, and sampling approach, we are unable to make direct statistical comparisons in the magnitude of responses between studies. Nevertheless, factors that may contribute to differences in vibriocidal titer between populations from different geographic regions include degree of “endemicity”, proportion with prior vaccination, and age distribution.

Oral cholera vaccine (OCV) campaigns can effectively increase immunity through both individual and herd-based immunity in high-risk areas, but the duration depends critically on population drivers such as vaccine coverage, waning vaccine efficacy, and net influx of susceptible people through population mobility [9]. Indeed, preventive OCV campaigns conducted in the study area in the years prior to the study period showed that absence and unawareness were the main reasons for non-vaccination [10]. Furthermore, our recent qualitative work on the social dynamics influencing cholera risk in this area identified community skepticism regarding efficacy of recent vaccination campaigns, and misinformation about the vaccine’s supposed dangers and side effects, which may have discouraged individuals vaccination uptake. While longitudinal serological data could help guide the timing of future vaccination campaigns, it is crucial to address the sociological factors that may contribute to distrust of the cholera vaccine and the humanitarian infrastructures responsible for its distribution among at-risk communities.

Our study had several limitations. Study participants were limited to individuals who could access cholera treatment centers. Due to low sample size, our study was not powered to evaluate the difference in antibody response among vaccinated and unvaccinated individuals. Although the vibriocidal response is the best non-mechanistic correlate of protection against *V. cholerae* [4][11], the vibriocidal titer alone has not demonstrated to be completely predictive of long-term immunity against clinical disease.

Nevertheless, we present the post-infection vibriocidal kinetics of a large cohort of culture-confirmed cholera patients in an endemic region of Africa and show evidence that younger children may have a faster decay rate than older persons. Our study begins to address the data gap in longitudinal antibody response studies outside of South Asia and suggests that more frequent boosting with vaccine may be needed for cholera-vulnerable communities like those in Goma, DRC.

## Data Availability

All data produced in the present study are available upon reasonable request to the authors

## Author contributions

*A*.*A*., *J*.*G*.*M*., *and D*.*T*.*L*. conceived and planned the experiments and acquired funding for the study. F.M. and M.A. recruited and enrolled patients and collected samples for the study. K.L.D. and C.N. carried out the laboratory assays. K.L.D., C.N., B.J.B. C.C., and C.S. performed data management and statistical analysis. K.L.D., C.N., and D.T.L. wrote the first draft of the manuscript. All authors reviewed and edited the manuscript.

## Acknowledgements

We acknowledge the contributions of DRC National Program for the Elimination of Cholera and Other Diarrheal Diseases (PNECHOL-DM); The North Kivu Provincial Health Division (DPS-NK); Appui Medical Integre aux Activites de Laboratoire (AMI-LABO); HEAL Africa.

## References

[1] S. Kanungo, A. S. Azman, T. Ramamurthy, J. Deen, and S. Dutta, “Cholera,” 2022. doi: 10.1016/S0140-6736(22)00330-0.

[2] World Health Organization, “Cholera. https://www.who.int/news-room/fact-sheets/detail/cholera, WHO.

[3] A. S. Azman, S. M. Moore, and J. Lessler, “Surveillance and the global fight against cholera: Setting priorities and tracking progress,” 2020. doi: 10.1016/j.vaccine.2019.06.037.

[4] A. S. Azman et al., “Estimating cholera incidence with cross-sectional serology,” Sci Transl Med, vol. 11, no. 480, 2019, doi: 10.1126/SCITRANSLMED.AAU6242.

[5] P. A. Harris, R. Taylor, R. Thielke, J. Payne, N. Gonzalez, and J. G. Conde, “Research electronic data capture (REDCap)-A metadata-driven methodology and workflow process for providing translational research informatics support,” J Biomed Inform, vol. 42, no. 2, 2009, doi: 10.1016/j.jbi.2008.08.010.

[6] A. S. Iyer et al., “Dried Blood Spots for Measuring Vibrio cholerae-specific Immune Responses,” PLoS Negl Trop Dis, vol. 12, no. 1, 2018, doi: 10.1371/journal.pntd.0006196.

[7] R. D. C. Team, “R: A language and Environment for statistical computing [Computer software].,” 2012.

[8] B. A. Muzembo, K. Kitahara, D. Mitra, A. Ohno, and S. I. Miyoshi, “Long-Term Kinetics of Serological Antibodies against Vibrio cholerae Following a Clinical Cholera Case: A Systematic Review and Meta-Analysis,” Int J Environ Res Public Health, vol. 19, no. 12, p. 7141, Jun. 2022, doi: 10.3390/IJERPH19127141/S1.

[9] C. M. Peak, A. L. Reilly, A. S. Azman, and C. O. Buckee, “Prolonging herd immunity to cholera via vaccination: Accounting for human mobility and waning vaccine effects,” PLoS Negl Trop Dis, vol. 12, no. 2, 2018, doi: 10.1371/journal.pntd.0006257.

[10] E. Briskin et al., “Oral cholera vaccine coverage in Goma, Democratic Republic of the Congo, 2022, following 2019–2020 targeted preventative mass campaigns,” Vaccine X, vol. 20, p. 100555, Oct. 2024, doi: 10.1016/j.jvacx.2024.100555.

[11] M. S. Son and R. K. Taylor, “Vibriocidal Assays to Determine the Antibody Titer of Patient Sera Samples,” Curr Protoc Microbiol, vol. 23, no. 1, pp. 6A.3.1-6A.3.9, Nov. 2011, doi: 10.1002/9780471729259.MC06A03S23.

